# Surgical candidacy and treatment uptake among women with cervical cancer at public referral hospitals in Kampala, Uganda

**DOI:** 10.1101/2019.12.31.19016279

**Authors:** Megan L Swanson, Miriam Nakalembe, Lee-may Chen, Stefanie M Ueda, Jane Namugga, Carol Nakisige, Megan J Huchko

## Abstract

**Purpose:** Cervical cancer is the most common malignancy among women in Uganda. Most present with advanced disease, when hysterectomy is not possible and cure is less likely. This study reports the proportion recommended for hysterectomy and associated factors, recommended treatments by stage, and treatment uptake.

**Methods:** We conducted a prospective study among patients seeking care for cervical cancer at public referral hospitals in Uganda. In-person surveys were followed by a phone call. Descriptive and multivariate statistical analyses examined associations between predictors and outcomes.

**Results:** Among 268 participants, 76% were diagnosed at an advanced stage (IIB-IVB). In total, 12% were recommended for hysterectomy. In adjusted analysis, living within 15 kilometers of Kampala (OR 3.10, 95% CI 1.20-8.03) and prior screening (OR 2.89, 95% CI 1.22-6.83) were significantly associated with surgical candidacy. Radiotherapy availability was not significantly associated with treatment recommendations for early-stage (IA-IIA) disease, but was associated with recommended treatment modality (chemo-radiation versus primary chemotherapy) for locally advanced stage (IIB-IIIB). Most (67%) had started treatment. No demographic or health factor, treatment recommendation, or radiation availability was associated with treatment initiation. Among those recommended for hysterectomy, 55% underwent surgery. Among those who had initiated treatment, 82% started the modality that was actually recommended.

**Conclusion:** Women presented to public referral centers in Kampala with mostly advanced-stage cervical cancer and few were recommended for surgery. Lack of access to radiation did not significantly increase the proportion of early-stage cancers recommended for hysterectomy.

## Introduction

Cervical cancer is the fourth most-common cancer among women worldwide: 570,000 new cases and 311,000 deaths estimated in 2018.^1^ Cervical cancer is over-represented in low- and middle-income countries (LMICs); the highest incidence and mortality rates are in Southern and Eastern Africa. In Uganda, cervical cancer is the most common malignancy and responsible for the greatest cancer-related mortality among women.^2^ While population-based cytologic screening has dramatically reduced incidence in high-income countries, incidence and mortality have risen in LMICs like Uganda,^3^ where less than 5% of women have ever been screened.^4^

In Uganda, most women with cervical cancer are diagnosed in an advanced stage,^5^ although poor record keeping affects the accuracy of estimates. The Kampala Cancer Registry lacked stage data for half of the 261 cervical cancer cases reported in a 3-year period; 73% of the remaining cases were Stage II+.^6^ As a result, 5-year overall survival for all cervical cancer cases is approximately 18%.^7^ Cervical cancer, especially early-stage, is potentially curable with surgery and/or radiation with or without chemotherapy, but multi-system obstacles often block access to treatment.^8^

Challenges to accessing surgery include limited specialty surgical training,^9,10^ few anesthesia providers,^11,12^ and a lack of sufficient banked blood.^13^ Access to radiation, gold-standard treatment for bulky early-stage disease and loco-regionally advanced cervical cancer, is limited in sub-Saharan Africa;^14^ Uganda is no exception.^15^ After the single functioning radiation machine was determined broken beyond repair in March 2016, Uganda was without radiotherapy until the new cobalt machine was inaugurated January 2018. During this time, patients had to travel to neighboring Kenya to access radiation. Chemotherapy is provided free of charge at government facilities, but shortages are frequent.^16^

Most care for cervical cancer in Uganda is provided at two specialty public hospitals: Mulago National Referral Hospital (MNRH), and the Uganda Cancer Institute (UCI). Clinical consultations are free of charge; there are nominal fees associated with radiotherapy and surgery. In response to the challenges obtaining radiotherapy 2016-2018, providers at UCI and MNRH developed a protocol for treating locally and regionally-advanced cervical cancer with neoadjuvant chemotherapy followed by possible radical hysterectomy, an evidence-based strategy supported by the American Society of Clinical Oncology (ASCO) Resource-stratified Treatment Guidelines.^17-20^

The ASCO Resource-stratified Treatment Guidelines also suggest a broader role for hysterectomy for cervical cancer in settings without access to radiation. While radical hysterectomy is a theoretical consideration for stage IA2 to IIA2, in maximally-resourced settings, chemoradiation is the preferred treatment modality for women with stage IB2 to IIA2 disease given equivalent outcomes with less morbidity.^21^ In settings without radiotherapy, primary treatment with hysterectomy may be considered as an alternative for women with stage IB2 to IIA2 disease.^17^

We sought to describe the presentation of cervical cancer at MNRH and UCI in Kampala, Uganda, specifically the proportion of patients recommended for hysterectomy (simple or radical) at the time of diagnosis, and factors associated with surgical candidacy. We also sought to describe primary treatment recommendations, utilization patterns and factors associated with successful treatment uptake, in the context of variably available treatments.

## Methods

From April 2017 through September 2018, we surveyed women over 18 years old presenting to care at either of two government-sponsored referral hospitals (MNRH and UCI) with a new diagnosis of cervical cancer and invited participation in baseline and follow-up surveys. This analysis is part of a larger study examining patterns of delay in accessing treatment.

Research assistants approached women in clinic and invited them to participate. Interested participants were screened for eligibility. Women >18 years old able to understand English or Luganda with a histopathologic diagnosis of primary cervical cancer and a clinical stage assigned by a gynecologist in the gynecologic oncology division at MNRH or UCI were eligible to participate. Participation was voluntary and participants were reimbursed in part for their travel expenses (20,000 UGX, approximately US$5). Individual informed consent was obtained.

Part one of the survey was administered in the clinic. Research assistants collected demographic and basic health information, including a reproductive health history from participants before collecting information about each individual’s journey to care. This quantitative survey was adapted from a validated survey instrument to measure time intervals and factors correlated with delay in accessing breast cancer care.^22^ Minor modifications were made in order to map the survey onto the Model of Pathways to Treatment, a theoretical model adapted from the Andersen Model for understanding and describing the process of obtaining diagnosis and treatment for cancer,^23-25^ and to make the questions specific to cervical cancer. Participants’ medical records were used to corroborate information on the histology, stage and grade of cervical tumors as well as the dates of the biopsy, staging exam and referral for treatment. Research assistants were in present in clinic and recruiting about 50% of the time during the study period.

Follow-up contact information was obtained. A telephone survey regarding treatment initiation was administered a minimum of four weeks and up to three months after the initial interview. Those who had not been able to access treatment were offered a follow-up visit and/or directions for follow-up in radiation oncology, medical oncology, or palliative care, as appropriate.

Survey data was captured on tablets using Open Data Kit (ODK) software (https://opendatakit.org).^26^ Data were uploaded daily and aggregated on a secure server.

This study was reviewed and approved by institutional review boards at MNRH, UCI and the University of California at San Francisco. The informed consent document and the survey were translated into Luganda; English is the official language of instruction at Makerere University College of Medical Sciences. Written informed consent documents were obtained from all participating survey responders.

To calculate our sample size, we assumed that if 20% of women with cervical cancer in Uganda were diagnosed with early-stage (I-IIA) cervical cancer, a total of 265 women would allow us to experimentally determine this proportion within 5% (confidence level 95%). As hysterectomy would be a theoretical treatment option for those with stage IA-IIA, this was also the theoretical proportion of surgical candidates, our primary outcome. We thus aimed to include at least 265 patients using consecutive sampling techniques.

Our primary outcome variable, surgical candidacy, was defined as a recommendation for hysterectomy (simple or radical) as primary treatment. While any patient with “early-stage” (IA to IIA) disease may have been considered for hysterectomy, we used the physician’s actual recommendation, after clinical assessment, to define “candidacy.” Any “late-stage” (IIB to IVB) patients recommended for surgery were dropped, as this would likely be a recommendation for a palliative surgery, not definitive primary therapy with curative intent. Treatment initiation, our other outcome variable, was defined by a participant self-reporting starting any treatment, including hospice.

To test for associations between categorical outcome variables (surgical candidacy and treatment initiation) and categorical explanatory variables, we performed chi-square tests and Fisher’s exact tests. For continuous explanatory predictors, we used two independent sample t-tests to compare sample means by outcome. We used logistic regression to explore the association of demographic and reproductive health variables with these outcomes. Factors theorized to be associated with the outcomes or those found to be significantly associated in bivariate analysis were considered for multivariate analysis. Because only those with early stage disease could be surgical candidates (see above), the variables were not independent; thus, we could not adjust analysis of surgical candidacy for stage. All data were analyzed using Stata version 14.0 (Stata Corporation, College Station, TX). P values less than 0.05 were considered statistically significant.

## Results

From April 2017 through September 2018, 332 women were screened and ultimately 268 participated (see Figure 1). Among participants, 233 (87%) initially presented with symptoms. Most women, (N=204, 76%) were diagnosed at an advanced stage (IIB to IVB) and these women, compared to those with early-stage (IA1 to IIA) disease (N=64, 24%), were more likely to present with symptoms including bleeding, discharge, pain or fatigue (OR 2.44, 95% CI 1.16-5.14) rather than through routine screening. About half of those with early-stage disease were recommended to undergo hysterectomy as primary treatment (N=33, 52% of those with early-stage disease, 12% of total).

**Figure 1:**
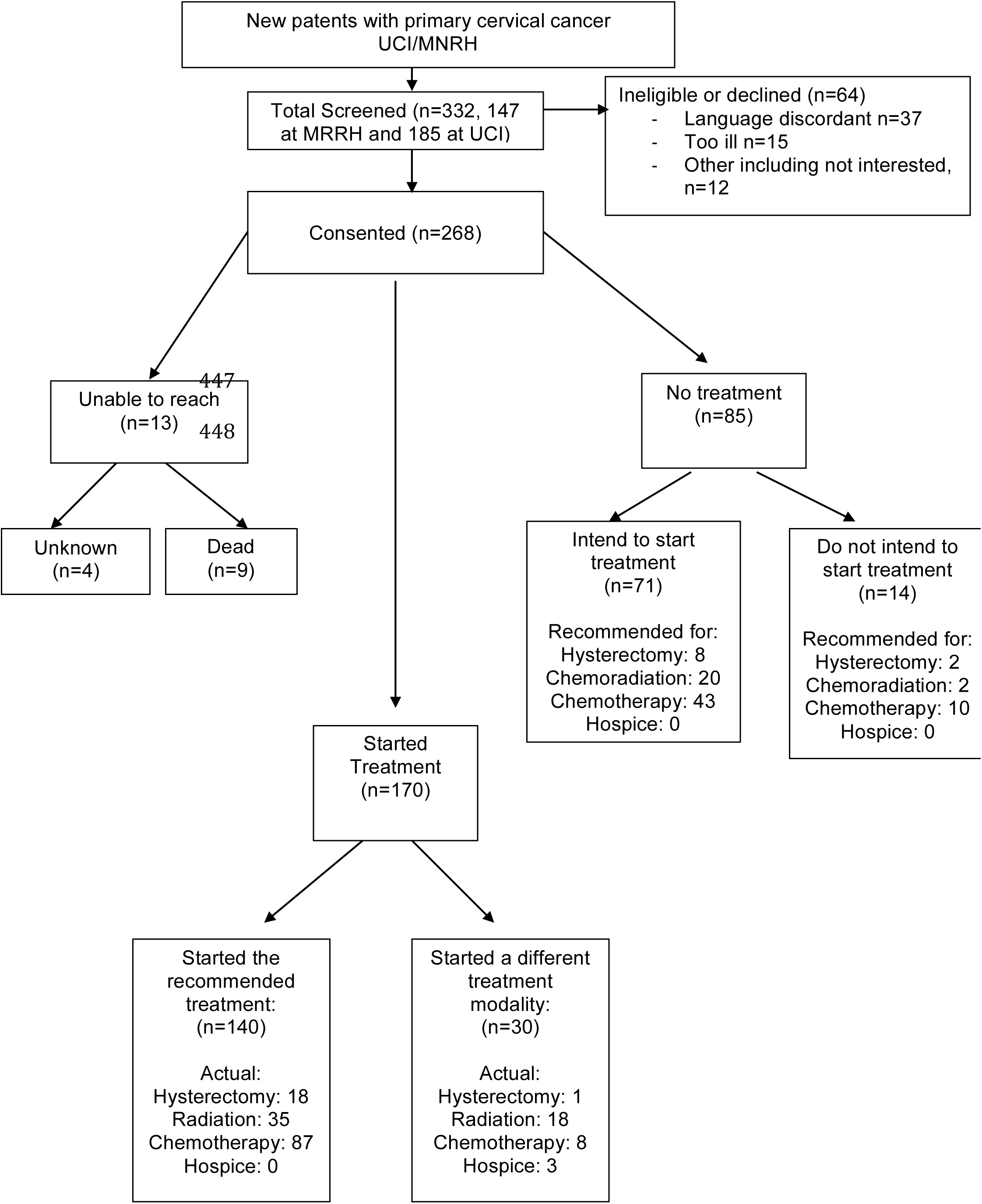
Study Flow Diagram. This flowchart depicts the treatment uptake patterns of study participants. It shows what treatments the patients who had NOT yet started treatment were recommended for by whether or not they planned to uptake treatment and the actual treatments started by women according to whether they started a recommended, versus other, treatment modality. Further details regarding recommendations by stage are available in Table 2 and Figure 2. Abbreviations: UCI, Uganda Cancer Institute; MNRH, Mulago National Referral Hospital.

In unadjusted analysis, living in or within 15 kilometers of Kampala (OR 3.38, 95% CI 1.60-7.13), current use of modern family planning (OR 3.98, 95% CI 1.26-12.49) and history of prior screening (OR 2.85, 95% CI 1.34-6.03) were associated with a recommendation for surgery. In multivariate analysis, living within 15 kilometers of Kampala (OR 3.10, 95% CI 1.20-8.03) and prior screening (OR 2.89, 95% CI 1.22-6.83) remained significantly associated with a recommendation for surgery as primary treatment (see Table 1).

**Table 1:** Characteristics of women diagnosed with cervical cancer by surgical candidacy. Surgical candidacy is defined as a recommendation for surgery (simple versus radical hysterectomy) after evaluation by a gynecologist at UCI or MNRH. We eliminated one participant with stage IIB disease who reported a recommendation for primary surgery as this was unlikely a recommendation for a curative hysterectomy (radical versus simple). Non-surgical candidates were recommended for primary treatment with chemoradiation, chemotherapy or hospice. The multivariate model was adjusted for age, education, distance, urban residence, marital status, parity, family planning use, history of prior screening, and HIV serostatus. Abbreviations: UCI, Uganda Cancer Institute; MNRH, Mulago National Referral Hospital; km, kilometers; HIV, Human Immunodeficiency Virus; CI, confidence interval; CA, cancer.

| Variable | Total<br>N | Surgical<br>candidates*<br>% | Not surgical<br>candidates**<br>% | Unadjusted Odds<br>Ratio of surgical<br>candidacy<br>(95% CI) | Adjusted***<br>Odds Ratio of<br>surgical<br>candidacy<br>(95% CI) |
| --- | --- | --- | --- | --- | --- |
| Total (N, %) | N=267 | N=33 (12%) | N=204 (88%) |  |  |
| Age, dichotomous |  |  |  |  |  |
| >= 50 years | 123 | 11 | 89 | 1.0 | 1.0 |
| < 50 years | 133 | 14 | 86 | 1.30 (0.62-2.72) | 0.94 (0.36-2.48) |
| Education |  |  |  |  |  |
| < primary | 110 | 9 | 91 | 1.0 | 1.0 |
| ≥ primary | 151 | 15 | 85 | 1.80 (0.82-3.95) | 1.40 (0.57-3.44) |
| Occupation |  |  |  |  |  |
| Industry/business | 92 | 12 | 88 | 1.0 | **** |
| Farming/domestic | 175 | 13 | 87 | 1.06 (0.49-2.29) | **** |
| Distance to MNRH/UCI |  |  |  |  |  |
| > 15m | 181 | 8 | 92 | 1.0 | 1.0 |
| ≤ 15km | 86 | 22 | 78 | 3.38 (1.60-7.13) | 3.10 (1.20-8.03) |
| Urban versus rural |  |  |  |  |  |
| Rural | 125 | 9 | 91 | 1.0 | 1.0 |
| Urban | 142 | 15 | 85 | 1.90 (0.88-4.09) | 1.15 (0.42-3.10) |
| Marital status |  |  |  |  |  |
| Single/divorced<br>/widowed | 146 | 10 | 90 | 1.0 | 1.0 |
| Married | 121 | 16 | 84 | 1.76 (0.84-3.67) | 1.26 (0.54-2.95) |
| Parity |  |  |  |  |  |
| ≤ 6 | 157 | 13 | 87 | 1.0 | 1.0 |
| > 6 | 110 | 12 | 88 | 0.92 (0.44-1.93) | 1.35 (0.51-3.51) |
| Family Planning |  |  |  |  |  |
| No method | 251 | 11 | 89 | 1.0 | 1.0 |
| Using a method | 15 | 33 | 67 | 3.98 (1.27-12.49) | 2.44 (0.61-9.76) |
| Prior Screening |  |  |  |  |  |
| No prior screening | 199 | 9 | 91 | 1.0 | 1.0 |
| History prior screening | 68 | 22 | 78 | 2.85 (1.34-6.03) | 2.89 (1.22-6.83) |
| HIV serostatus |  |  |  |  |  |
| HIV - | 179 | 13 | 87 | 1.0 | 1.0 |
| HIV + | 82 | 11 | 89 | 0.84 (0.37-1.90) | 0.52 (0.18-1.45) |
| Previously heard of cervical cancer |  |  |  |  |  |
| Never heard | 58 | 10 | 90 | 1.0 | **** |
| Heard of cervix CA | 209 | 13 | 87 | 1.29 (0.50-3.28) | **** |
| Know friend/family with cervical cancer |  |  |  |  |  |
| Don't know | 214 | 12 | 88 | 1.0 | **** |
| Know friend/family | 53 | 13 | 87 | 1.10 (0.45-2.69) | **** |

\* As recommended by specialists at UCI/MNRH; eliminating one women stage IIB who reported a recommendation for surgery
\*\*\*\* Not included in the multivariate model

Recommended treatments varied by stage, as expected. We hypothesized that recommendations may also vary by availability of radiation therapy (see Table 2). Almost two-thirds of the participants (N=167, 62%) presented for care at a time when no in-country external beam radiation was available, compared to 101 participants (38%) who received treatment recommendations after the new machine was commissioned.

**Table 2:** Treatment recommendations by stage and availability of radiation. In-country radiation availability was determined by date of diagnosis and whether the new cobalt machine was functional at that time. All patients recommended for external beam radiation were also recommended to have concurrent chemo-sensitization, typically with weekly cisplatin 40mg per square-meter concurrent with external beam pelvic radiation to a total of 4500cGy in 25 fractions. Participants recommended for chemotherapy may have been either a neoadjuvant approach or a palliative approach, especially for metastatic disease. Chemotherapy would typically be doublet therapy with cisplatin 75mg per square-meter and paclitaxel 135-175mg per square-meter. Abbreviations: RT, radiation; mg, milligrams.

| Stage at Presentation | Radiation Availability | Surgery | Chemoradiation* | Chemotherapy** | Hospice |
| --- | --- | --- | --- | --- | --- |
| **IA1-IB1**<br>N=35 | No RT available (N=23) | 21 (92%) | 1 (4%) | 1 (4%) | 0 |
|  | RT available (N=12) | 11 (92%) | 0 | 1 (8%) | 0 |
| **IB2-IIA**<br>N=29 | No RT available (N=16) | 1 (6%) | 3 (19%) | 12 (75%) | 0 |
|  | RT available (N=13) | 0 | 6 (46%) | 7 (54%) | 0 |
| **IIB-IIIB**<br>N=190 | No RT available (N=119) | 1 (1%) | 11 (9%) | 107 (90%) | 0 |
|  | RT available (N=71) | 0 | 45 (63%) | 26 (37%) | 0 |
| **IVA-IVB**<br>N=14 | No RT available (N=9) | 0 | 0 | 8 (89%) | 1 (11%) |
|  | RT available (N=5) | 0 | 2 (40%) | 2 (40%) | 1 (20%) |

\*All patients recommended for external beam radiation were also recommended to have concurrent weekly chemo-sensitization with cisplatin 40mg per square-meter concurrent with external beam pelvic radiation, 4500cGy in 25 fractions
\*\*Chemotherapy recommended as primary modality, either in neoadjuvant setting (stage IB1 – IIB) or for distant metastases (stage IV). Doublet therapy with cisplatin 75mg per square-meter and paclitaxel 135-175mg per square-meter every three weeks was the standard recommended regimen.

Most women with stage IA1 to IB1 disease (N=35) were recommended for surgery regardless of availability of in-country radiation (N=32, 92%). Among women with stage IB2-IIA disease (N=29), only one was recommended to undergo hysterectomy and this was when the radiation machine was broken down. Surprisingly, chemotherapy (likely neoadjuvant) was recommended as the primary treatment to most of those with IB2-IIA disease, regardless of availability of radiation (19/29, 66% overall; 75% when radiation machine broken down versus 54% when machine working, OR 2.57, 95% CI 0.53-12.38).

However, for women with locally advanced disease (stage IIB-IIIB), absence of in-country radiation was associated with significantly higher odds of a recommendation for chemotherapy (OR 15.43, 95% CI 7.16-33.25) and lower odds of a recommendation for chemoradiation (OR 0.06, 95% CI 0.03-0.13). Few women (N=14) had stage IV disease and none of these women were recommended for radiation when the machine was down. Only two participants, both stage IV, were recommended to start hospice.

We obtained follow-up information for 264 of the 268 participants. Nine women died before starting treatment. Among the 255 living women reached for phone interview, 170 (67%) had initiated some treatment. No demographic or health or cancer-related variables (including treatment recommendations and radiation availability) were associated with treatment initiation in univariate analysis (see Table 3). Given lack of association, multivariate analysis was not performed.

**Table 3:** Characteristics of participants by whether or not they had started treatment. “Treatment” refers to any treatment (surgery, chemoradiation, chemotherapy, hospice), whether it was the recommended treatment or a different modality. The table excludes the nine women who died by the time their contacts were reached for follow-up (four had been recommended for chemotherapy, four for chemoradiation and one for hospice) and the four who were unable to be reached. A multivariate analysis was not performed given lack of association in unadjusted analysis. Abbreviations: UCI, Uganda Cancer Institute; MNRH, Mulago National Referral Hospital; km, kilometers; RT, radiation; HIV, Human Immunodeficiency Virus; CI, confidence interval; CA, cancer.

| Variable | Total<br>N | No treatment<br>% | Started treatment<br>% | Unadjusted Odds<br>Ratio of Treatment<br>initiation<br>(95% CI) |
| --- | --- | --- | --- | --- |
| Total (N, %)* | N=255 | N= 85 (33%) | N= 170 (67%) |  |
| Age, dichotomous |  |  |  |  |
| >= 50 years | 115 | 34 | 66 | 1.0 |
| < 50 years | 129 | 32 | 68 | 1.10 (0.64-1.88) |
| Education |  |  |  |  |
| < primary | 106 | 28 | 72 | 1.0 |
| ≥ primary | 143 | 36 | 64 | 0.69 (0.40-1.19) |
| Occupation |  |  |  |  |
| Industry/business | 90 | 31 | 69 | 1.0 |
| Farming/domestic | 165 | 35 | 65 | 0.86 (0.49-1.48) |
| Distance from MNRH/UCI |  |  |  |  |
| ≤ 15km | 84 | 27 | 73 | 1.51 (0.85-2.67) |
| > 15m | 171 | 36 | 64 | 1.0 |
| Urban versus rural |  |  |  |  |
| Rural | 118 | 37 | 63 | 1.0 |
| Urban | 137 | 30 | 70 | 1.39 (0.83-2.35) |
| Marital status |  |  |  |  |
| Single/divorced/widowed | 138 | 37 | 63 | 1.0 |
| Married | 117 | 29 | 71 | 1.43 (0.84-2.42) |
| Parity |  |  |  |  |
| ≤ 6 | 148 | 38 | 63 | 1.0 |
| > 6 | 107 | 27 | 73 | 1.64 (0.95-2.81) |
| Family Planning |  |  |  |  |
| No method | 239 | 33 | 67 | 1.0 |
| Using a method | 15 | 33 | 67 | 0.99 (0.33-2.99) |
| Prior Screening |  |  |  |  |
| None prior | 189 | 33 | 67 | 1.0 |
| History prior screening | 66 | 33 | 67 | 1.0 (0.55-1.81) |
| HIV serostatus |  |  |  |  |
| HIV - | 170 | 34 | 66 | 1.0 |
| HIV + | 79 | 30 | 70 | 1.16 (0.65-2.06) |
| Early stage at diagnosis |  |  |  |  |
| Stage IIB-IVA | 191 | 34 | 66 | 1.0 |
| Stage IA1-IIA | 64 | 33 | 67 | 1.03 (0.57-1.88) |
| Previously heard of cervical cancer |  |  |  |  |
| Never heard | 57 | 33 | 67 | 1.0 |
| Heard of cervix CA | 198 | 33 | 67 | 1.0 (0.54-1.87) |
| Know friend/family with cervical cancer |  |  |  |  |
| Don't know | 205 | 32 | 68 | 1.0 |
| Know friend/family | 50 | 40 | 60 | 0.70 (0.37-1.32) |
| Radiation machine working at time of diagnosis |  |  |  |  |
| RT not available | 159 | 32 | 68 | 1.0 |
| RT available | 96 | 35 | 65 | 0.86 (0.50-1.47) |
| Recommended treatment modality |  |  |  |  |
| Surgery | 33 | 30 | 70 | 1.0 |
| Chemotherapy | 159 | 33 | 67 | 0.83 (0.37-1.87) |
| Chemoradiation | 61 | 36 | 64 | 0.74 (0.30-1.82) |

\* Excluding 9 women who died by the time their contacts were reached for follow-up (4 had been recommended for chemotherapy, 4 for chemoradiation and 1 for hospice) and 4 who were unable to be reached.

Of the 33 women recommended to undergo hysterectomy, 55% actually had surgery, another 15% started other treatment modalities and the remaining 30% had not started any treatment (see Fig 2; of note, one woman stage IB1 who was recommended for chemotherapy ultimately had a hysterectomy). Among the 170 women who started treatment, most (N=140, 82%) of them started the treatment modality that was actually recommended. Only young age (< 50 years) was associated with higher odds of starting the recommended treatment (OR 1.84, 95% CI 1.11-3.07); there was no association with any other demographic or health or cancer-related factors (including stage, treatment recommendations, and radiation availability).

**Figure 2:**
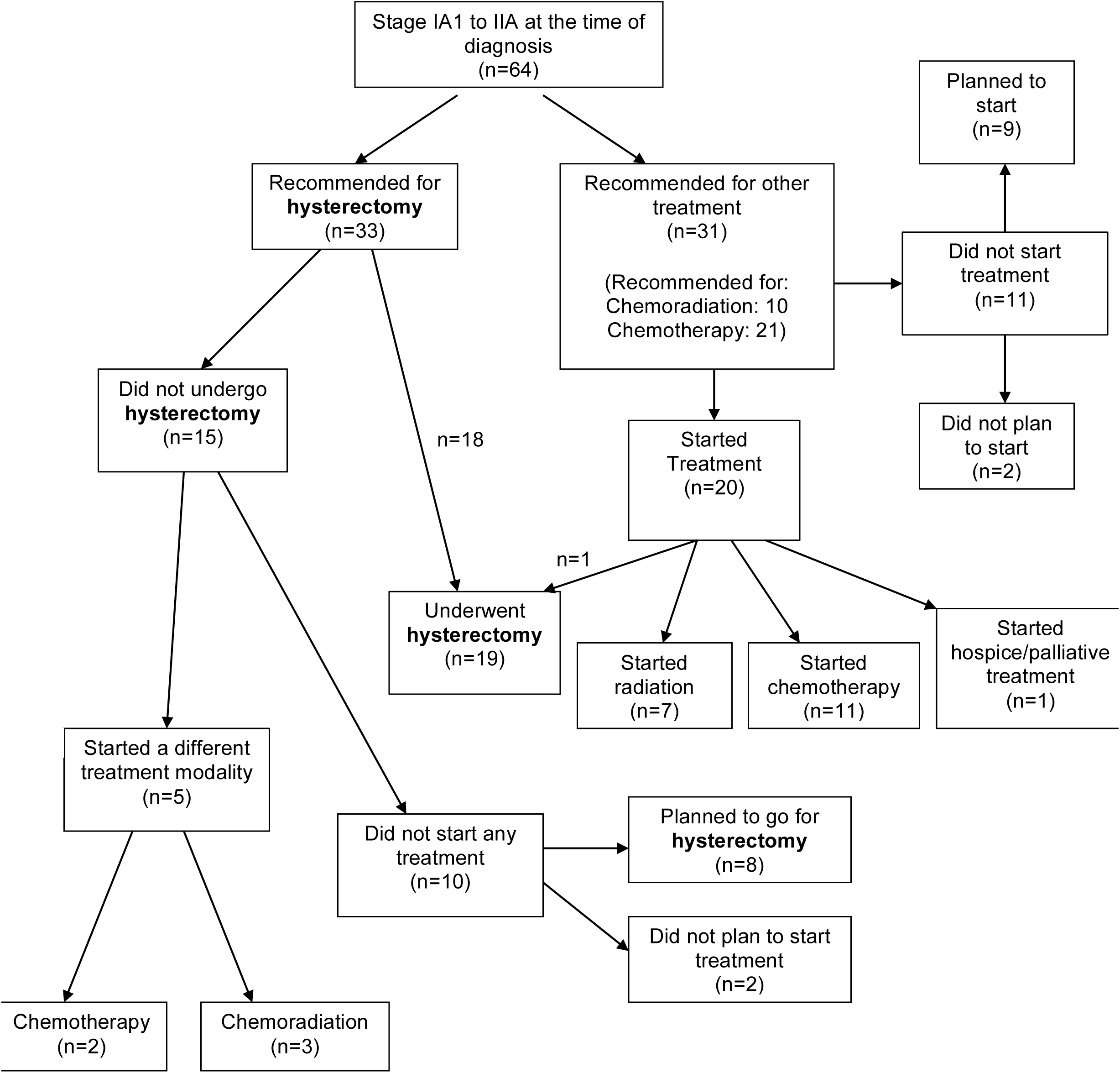
Treatment recommendations and uptake for women with early-stage disease, especially those recommended for hysterectomy. The flowchart shows whether or not treatment was initiated by recommendation for hysterectomy or other treatment. The chart goes on to show the treatments actually started, whether or not these were the recommended treatments. The chart also shows, for those who had NOT yet started treatment, whether or not they planned to undergo treatment, by initial recommendation. The chart shows that a total of 19 women with early-stage disease ultimately underwent hysterectomy, though only 18 of these women had initially been recommended for hysterectomy. No abbreviations.

Of the 85 participants who had not started treatment, 71 (84%) planned to start. The most commonly reported reasons for delay were financial constraints (69%) and long wait times (30%). For the subset of these women recommended for hysterectomy (N=8), the most common reasons for delay were the same: financial constraints (50%) and perceived long wait (37.5%). For the 14 participants not planning to start any treatment or hospice, the most common reasons were pursuing alternative or traditional methods (57%) and/or financial constraints (50%). Two of these 14 had been recommended for hysterectomy, one cited “a long queue” and the other distrust of treatment recommendation as the reason for not pursuing treatment.

## Discussion

In this cohort of women with cervical cancer presenting at referral centers in Kampala, Uganda, three-quarters presented at an advanced stage (IIB-IVB), and only 12% were recommended to undergo hysterectomy. Overall, treatment recommendations were in-line with the ASCO Resource-Stratified Clinical Practice Guideline.^17^ The significance of the availability of radiation in Uganda on treatment recommendations seemed to vary based on stage, though the study was underpowered to assess these effects by subgroup.

This study adds to the epidemiology of cervical cancer in the region and is the first study in the region to report on proportion of newly diagnosed cervical cancer patients recommended for hysterectomy. The proportion of participants presenting at late stage (IIB-IVB), 76%, is similar to 20 year-old estimates from the Kamala Cancer Registry,^6^ as well as to more recent regional estimates from Northern Uganda,^5^ Ghana,^27^ and Rwanda.^28^ Given the lack of national screening and nascent HPV vaccination efforts, it is unsurprising that the proportion of late stage disease is unchanged over the last two decades in Uganda.

As expected, women with early-stage disease were more likely to be assessed as surgical candidates. Although radical hysterectomy is a theoretical consideration for stage IB2-IIA disease, chemoradiation, when available, is preferred. We expected more of these women to be recommended for primary surgery when the radiation machine was out of commission. In this subset (stage IB2-IIA), there was a non-significant trend to increased recommendation for radical hysterectomy during the time without in-country radiation, though the study was underpowered to assess this impact. Although large tumors (>4cm) are not necessarily associated with increased complications among experienced surgeons,^21,29^ newly trained providers with limited resources including a lack of equipment and cross-matched blood may not be eager to offer radical hysterectomy to stage IB2-IIA patients.

Surprisingly, the majority of the women stage IB2-IIA were recommended for chemotherapy (likely neoadjuvant), regardless of the availability of radiation. As chemoradiation is the preferred modality, we would have expected more of these women to be referred for radiation when the new machine was operational. Availability is not synonymous with accessibility; prior to discontinuation, the old machine, despite running 20 hours per day, was meeting just 2.6% of the nation’s indicated radiation treatments.^14,15^ Alternatively, it is possible a perception of success resulted in a continuation of the practice pattern of neoadjuvant chemotherapy. These data are forthcoming.

Two-thirds of women in the current study had initiated treatment; neither stage, recommended treatment, nor radiation availability predicted treatment initiation. Disappointingly, only 55% of the women recommended for curative hysterectomy were able to undergo surgery. Women unable to access hysterectomy reported the same challenges as women recommended for chemoradiation or chemotherapy.

There are comparable regional data on radiation acquisition. A survey from rural Rwanda reported that 80% of the cervical cancer patients at the Butaro Cancer Center of Excellence (BCCOE) who were referred to UCI for chemoradiation (there is no available radiation in Rwanda) were able to access treatment.^28^ This high proportion of treatment acquisition contrasts our findings that women were generally unable to access out-of-country radiation. As expected, in our study, women with locally advanced disease (stage IIB-IIIB) were less likely to be recommended for standard-of-care chemoradiation when there was no available in-country radiation. For those recommended to undergo chemoradiation in Kenya, when the radiation machine in Uganda was down (N=15), only two of them reported actually starting chemoradiation, whereas when the Kampala-based machine was working, two-thirds of women were able to access this treatment.

The most commonly cited reason for failing to access treatment, including curative hysterectomy, financial hardship, is difficult for the under-funded public health sector to modify. Clinical consultations are free of charge and there are nominal fees associated with radiation, surgery, labs and imaging. The second-most cited reason, long wait times, implies an unmet need for treatment, including radical hysterectomies. As the burden of cancer shifts to LMICs, a re-calibrated response from the international community is necessary to substantially increase funding for capacity building and training opportunities for local clinicians as well as for sufficient equipment and supplies to enable oncologic surgery, radiation and chemotherapy.^30^

Generally speaking, most participants had previously heard of cervical cancer, similar to reports from Northern Uganda,^31^ but neither knowledge nor knowing someone with cervical cancer was associated with surgical candidacy or successful treatment initiation. Thus, education efforts designed to increase awareness of cervical cancer would not likely increase the proportion of women diagnosed at an early stage in absence of screening and referral services.

This study highlights the prevalence of late presentation to cervical cancer care and suggests that distance from treatment facilities and general poor access to care (as evidenced by lack of prior screening) affect women’s eligibility for curative surgery. These risks are difficult to modify, but suggest that either disseminating specialty cancer care beyond large urban centers, as the Rwandan Ministry of Health did with the BCCOE, or strengthening the referral system between rural health centers and the referral hospitals may improve timeliness to care.

While this study provides a snapshot of cervical cancer epidemiology, treatment recommendations and uptake, the population is a convenience sample of women who were able to access care at referral centers, limiting generalizability. The study population likely represents a conservative estimate of those at a late stage at time of presentation and likely over-estimates the proportion of women nationally who may be surgical candidates at the time of diagnosis. The myriad challenges associated with providing cancer treatment including lack of available operating theaters, surgical equipment, anesthesiologists, and trained surgeons for early-stage disease as well as a lack of radiation, chemotherapy, and radiation and medical oncologists for late-stage disease, underscore the need to expand prevention and screening opportunities in Uganda.

Further research is needed to assess when and why women experience delay in accessing care. Additionally, hospice and palliative care, though widely available in Uganda and subsidized, seemed to be poorly utilized. Given the substantial proportion of women presenting at late stage, the nine women who died before accessing the recommended treatment as well as the significant proportion of women who were not able to access any treatment, hospice is likely an appropriate treatment strategy for many women. Future research is needed to understand barriers specific to uptake of palliative care and hospice.

While decreasing the incidence of cervical cancer will only be possible by expanding vaccination and screening opportunities, efforts to improve earlier detection and diagnosis, expand accessibility of surgery, radiation and chemotherapy, and integrate palliative care into standard treatment are essential to decrease mortality, morbidity and suffering among women with cervical cancer.

## Data Availability

Raw data is kept on a secure server at the Infectious Diseases Institute

## Acknowledgments

to data collection team: Najjemba Irene, Najjemba Beatrice, Nakadu Laurant, Nankya Esther

## Author Contributions

Conception and design: Swanson, Nakalembe, Chen, Huchko

Administrative support: Nakalembe, Namugga, Nakisige

Provision of study materials or patients: Nakalembe, Namugga, Nakisige

Collection and assembly of data: Swanson, Nakalembe

Data analysis and interpretation: Swanson, Huchko

Manuscript writing: All

Final approval of manuscript: All

Accountable for all aspects of this work: All

## References

1. Bray F, Ferlay J, Soerjomataram I, et al : Global cancer statistics 2018: GLOBOCAN estimates of incidence and mortality worldwide for 36 cancers in 185 countries. CA Cancer J Clin 68:394–424, 2018

2. Bruni L B-RL, Albero G, Aldea M, Serrano B, Valencia S, Brotons M, Mena M, Cosano R, Muñoz J, Bosch FX, de Sanjosé S, Castellsagué X: Human Papillomavirus and Related Diseases in Uganda. Summary Report 2016-02-26, ICO Information Centre on HPV and Cancer (HPV Information Centre), 2016

3. Wabinga HR, Nambooze S, Amulen PM, et al : Trends in the incidence of cancer in Kampala, Uganda 1991-2010. Int J Cancer 135:432–9, 2014

4. Ndejjo R, Mukama T, Musabyimana A, et al : Uptake of Cervical Cancer Screening and Associated Factors among Women in Rural Uganda: A Cross Sectional Study. PLoS One 11:e0149696, 2016

5. Mwaka AD, Garimoi CO, Were EM, et al : Social, demographic and healthcare factors associated with stage at diagnosis of cervical cancer: cross-sectional study in a tertiary hospital in Northern Uganda. BMJ Open 6:e007690, 2016

6. Wabinga H, Ramanakumar AV, Banura C, et al : Survival of cervix cancer patients in Kampala, Uganda: 1995-1997. Br J Cancer 89:65–9, 2003

7. Gondos A, Brenner H, Wabinga H, et al : Cancer survival in Kampala, Uganda. Br J Cancer 92:1808–12, 2005

8. Sullivan R, Alatise OI, Anderson BO, et al : Global cancer surgery: delivering safe, affordable, and timely cancer surgery. Lancet Oncol 16:1193–224, 2015

9. Johnston C, Ng JS, Manchanda R, et al : Variations in gynecologic oncology training in low (LIC) and middle income (MIC) countries (LMICs): Common efforts and challenges. Gynecologic Oncology Reports 20:9–14, 2017

10. Meara JG, Leather AJ, Hagander L, et al : Global Surgery 2030: evidence and solutions for achieving health, welfare, and economic development. Int J Obstet Anesth 25:75–8, 2016

11. Dubowitz G, Detlefs S, McQueen KA: Global anesthesia workforce crisis: a preliminary survey revealing shortages contributing to undesirable outcomes and unsafe practices. World J Surg 34:438–44, 2010

12. Epiu I, Tindimwebwa JV, Mijumbi C, et al : Challenges of Anesthesia in Low-and Middle-Income Countries: A Cross-Sectional Survey of Access to Safe Obstetric Anesthesia in East Africa. Anesth Analg 124:290–299, 2017

13. Lund TC, Hume H, Allain JP, et al : The blood supply in Sub-Saharan Africa: needs, challenges, and solutions. Transfus Apher Sci 49:416–21, 2013

14. Abdel-Wahab M, Bourque JM, Pynda Y, et al : Status of radiotherapy resources in Africa: an International Atomic Energy Agency analysis. Lancet Oncol 14:e168–75, 2013

15. Datta NR, Samiei M, Bodis S: Radiation therapy infrastructure and human resources in low-and middle-income countries: present status and projections for 2020. Int J Radiat Oncol Biol Phys 89:448–57, 2014

16. Barr R, Robertson J: Access to Cytotoxic Medicines by Children With Cancer: A Focus on Low and Middle Income Countries. Pediatr Blood Cancer 63:287–91, 2016

17. Chuang LT, Temin S, Camacho R, et al : Management and Care of Women With Invasive Cervical Cancer: American Society of Clinical Oncology Resource-Stratified Clinical Practice Guideline. Journal of Global Oncology 2:311–340, 2016

18. Benedetti-Panici P, Greggi S, Colombo A, et al : Neoadjuvant chemotherapy and radical surgery versus exclusive radiotherapy in locally advanced squamous cell cervical cancer: results from the Italian multicenter randomized study. J Clin Oncol 20:179–88, 2002

19. Chang TC, Lai CH, Hong JH, et al : Randomized trial of neoadjuvant cisplatin, vincristine, bleomycin, and radical hysterectomy versus radiation therapy for bulky stage IB and IIA cervical cancer. J Clin Oncol 18:1740–7, 2000

20. Kokka F, Bryant A, Brockbank E, et al : Hysterectomy with radiotherapy or chemotherapy or both for women with locally advanced cervical cancer. Cochrane Database Syst Rev:Cd010260, 2015

21. Landoni F, Maneo A, Colombo A, et al : Randomised study of radical surgery versus radiotherapy for stage Ib-IIa cervical cancer. Lancet 350:535–40, 1997

22. Unger-Saldana K, Pelaez-Ballestas I, Infante-Castaneda C: Development and validation of a questionnaire to assess delay in treatment for breast cancer. BMC Cancer 12:626, 2012

23. Scott SE, Walter FM, Webster A, et al : The model of pathways to treatment: conceptualization and integration with existing theory. Br J Health Psychol 18:45–65, 2013

24. Walter F, Webster A, Scott S, et al : The Andersen Model of Total Patient Delay: a systematic review of its application in cancer diagnosis. J Health Serv Res Policy 17:110–8, 2012

25. Andersen BL, Cacioppo JT: Delay in seeking a cancer diagnosis: delay stages and psychophysiological comparison processes. Br J Soc Psychol 34 (Pt 1):33–52, 1995

26. Hartung C, Lerer A, Anokwa Y, et al : Open data kit: tools to build information services for developing regions. Presented at the Proceedings of the 4th ACM/IEEE International Conference on Information and Communication Technologies and Development, London, United Kingdom, 2010

27. Dunyo P, Effah K, Udofia EA: Factors associated with late presentation of cervical cancer cases at a district hospital: a retrospective study. BMC Public Health 18:1156, 2018

28. Park PH, Davey S, Fehr AE, et al : Patient Characteristics, Early Outcomes, and Implementation Lessons of Cervical Cancer Treatment Services in Rural Rwanda. J Glob Oncol 4:1–11, 2018

29. Machida H, Matsuo K, Furusawa A, et al : Profile of treatment-related complications in women with clinical stage IB-IIB cervical cancer: A nationwide cohort study in Japan. PLoS One 14:e0210125, 2019

30. Swanson M, Ueda S, Chen L-m, et al : Evidence-based improvisation: Facing the challenges of cervical cancer care in Uganda. Gynecologic Oncology Reports 24:30–35, 2018

31. Mwaka AD, Orach CG, Were EM, et al : Awareness of cervical cancer risk factors and symptoms: cross-sectional community survey in post-conflict northern Uganda. Health Expect, 2015

